# Data Diversity vs. Model Complexity in the Prediction of Pediatric Bipolar Disorder: Evidence from Academic and Community Clinical Samples

**DOI:** 10.64898/2026.03.26.26349447

**Authors:** Zhuoyu Shi, Eric A. Youngstrom, Yinuo Liu, Jennifer K. Youngstrom, Robert L. Findling

## Abstract

Pediatric bipolar disorder is challenging to diagnose accurately due to symptom heterogeneity. More standardized and data-driven approaches are needed to enhance diagnostic reliability. We evaluated a clinical decision tool (nomogram), statistical methods (logistic regression, LASSO), machine learning (support vector machine, random forest, k-nearest neighbors, extreme gradient boosting), and deep learning model (multilayer perceptron) for pediatric bipolar disorder prediction across two datasets collected in academic (*N*=550) and community (*N*=511) clinical settings. We compared three modeling strategies: cross-dataset validation, cross-dataset with interaction terms, and mixed-dataset. We assessed model performance using discrimination ability, calibration, and predictor importance ranking.

In the baseline cross-dataset approach, all models showed good internal discrimination in the academic dataset; but external discrimination in the community dataset substantially declined. Interaction-enhanced models slightly improved internal discrimination but not external performance or calibration. Recalibration prominently improved cross-dataset calibration without compromising discrimination, indicating that transportability problems were largely driven by probability scaling. Models trained on mixed datasets exhibited much stronger external discrimination and calibration. Across models and training strategies, family risk and PGBI-10M were consistently ranked as the most important predictors.

Predictive models for pediatric bipolar disorder showed strong internal performance but limited cross-setting generalizability due to dataset shift and miscalibration. Increasing model complexity did not improve external performance, whereas training on pooled data substantially improved both discrimination and calibration. Findings suggest that sampling diversity, rather than model complexity, is more valuable for developing clinically useful and generalizable psychiatric prediction models, underscoring the importance of open and collaborative datasets.

## Introduction

Pediatric bipolar disorder (PBD) is one of the most challenging psychiatric conditions to accurately diagnose, especially when initial manifestations occur in children and adolescents (Jensen-Doss et al., 2014; Youngstrom et al., 2008). Characterized by significant fluctuations in mood, energy, and activity, PBD can substantially impair children’s daily functioning and development (Keenan-Miller and Miklowitz, 2011). Despite its clinical significance, reliable and timely diagnosis remains challenging. Compared to adults, pediatric presentations are often more heterogeneous and involve complex and rapid changes of mood states, making the traditional diagnostic criteria less straightforward to apply (Goldstein and Birmaher, 2012). Moreover, symptoms frequently overlap with other psychiatric conditions, particularly attention-deficit/hyperactivity disorder and anxiety disorder, contributing to frequent misdiagnosis and diagnostic delays that may extend for several years (Post et al., 2010; Singh and Rajput, 2006). Such delays are associated with worse clinical outcomes, including increased recurrence, comorbidity, and treatment resistance (Goldstein et al., 2017; McIntyre et al., 2020; Post et al., 2010). These challenges underscore the need for more reliable and objective approaches to support early and accurate diagnosis.

Accurate diagnosis is fundamental for all subsequent clinical decision-making, guiding treatment selection and influencing long-term outcomes. However, current diagnostic practices are often subject to variability and bias (Mokros et al., 2018). In clinical settings, unstructured clinical interviews remain widely used (Jones, 2010), relying on clinicians’ experience even though experience without feedback shows little relationship with accuracy (Spengler et al., 2009). While this approach can sometimes efficiently decompose complex situations into actionable diagnoses, it may lead to inconsistent diagnostic decisions (Jenkins and Youngstrom, 2016), including overdiagnosis of some conditions and underestimation of others (Jensen-Doss et al., 2014). Moreover, contextual factors, such as the order in which a patient’s symptoms are reported to clinicians, can further influence diagnostic outcomes (Cwik and Margraf, 2017). Therefore, more standardized and data-driven approaches are needed to enhance diagnostic reliability.

Advances in statistical and computational methods have increasingly enabled the development of predictive models to assist clinical decision-making. Prior work has shown that simple algorithms can perform comparably to, or even outperform, experienced clinicians across a range of mental health prediction tasks, including psychiatric diagnosis, substance use classification, suicide risk assessment, and violence risk prediction (Ægisdóttir et al., 2006). A wide range of approaches, including clinical decision tools, statistical models, machine learning, and deep learning models, have been proposed to predict the risk of psychiatric disorders and enhance diagnostic accuracy. Early studies focused on structured clinical tools and nomogram-based approaches derived from symptom scales and family history (Youngstrom et al., 2008; Youngstrom et al., 2018), while statistical models such as logistic regression and penalized approaches like LASSO have been used to identify key predictors (Harrell, 2001; Shen et al., 2024; Youngstrom et al., 2018). More recently, machine learning (ML) and deep learning (DL) approaches have become increasingly popular due to their ability to model complex, high-dimensional data (Bzdok and Meyer-Lindenberg, 2018; Shatte et al., 2019; Shi et al., 2024). These artificial intelligence models often report strong performance, with high accuracy in validation on their own training and testing datasets (Jan et al., 2021; Librenza-Garcia et al., 2017). However, their performance in real-world clinical settings remains uncertain.

A major limitation of existing models is their limited generalizability across clinical settings. This limitation is often driven by dataset shift, arising from differences in patient populations, assessment procedures, and clinical severity (Beam and Kohane, 2018; Konig et al., 2007; Sahiner et al., 2023). As a result, models trained in one setting may not transfer well to another, particularly when clinical samples differ in symptom presentation, referral patterns, or sociodemographic composition. In addition to reduced classification ability, models frequently exhibit poor calibration when applied to new populations, which may compromise their clinical utility (Steyerberg et al., 2010; Van Calster et al., 2019). Calibration reflects the agreement between predicted probabilities and observed outcomes and is particularly important in clinical contexts where risk estimates derived from these models may assist in diagnosis. Miscalibration can lead to unreliable risk estimation and inconsistent clinical decisions, including potential overdiagnosis or missed cases.

In this study, we systematically evaluated the performance and generalizability of multiple prediction models for pediatric bipolar disorder across two datasets with a shared diagnostic framework, with one from an academic clinic and one from a community clinic. We compared three sampling and modeling strategies, including cross-dataset, cross-dataset with interaction-enhanced, and mixed dataset approaches, across a spectrum of models ranging from clinical decision tools recommended by evidence-based medicine and evidence-based assessment to statistical models, machine learning, and deep learning techniques. A Bayesian approach using a probability nomogram or calculator to combine prior probability with likelihood ratios based on a few key clinical findings produces substantial improvements in risk estimates and diagnostic accuracy, but it also represents an upper bound in terms of the limits of training and human workforce implementation. More complex statistical models, machine learning, and deep learning all would require delegating more of the evaluation process from human decision-making to a hybrid approach. We compared model performance in terms of discrimination, calibration, and predictor importance. We aimed to inform the choice of best approach for translating predictive models into real-world psychiatric practice.

## Methods

### Participants

The participants were youths aged 5 to 18 years. The participants and their caregivers were recruited from outpatient mental health centers. Recruitment was conducted at University Hospitals/Case Western Reserve University and several community-based clinics operating under Applewood Centers in Cleveland, Ohio. The study’s protocols were approved by the Institutional Review Boards at both institutions. All youths and caregivers were fluent in English. Youths diagnosed with pervasive developmental disorders or cognitive impairments were not included in the study. All families involved received compensation for their contribution to the research.

Two datasets were collected under different referral patterns. The academic dataset (*N*=550) was collected at a clinic located within a university’s psychiatry department (Findling et al., 2005). These families were referred to the clinic from external sources or within the department for concerns related to youth mood symptoms. The community dataset (*N*=511) was a randomly selected subgroup from mental health and behavioral services. This group participated in both the standard intake process and a comprehensive enhanced research interview (Youngstrom et al., 2005). A summary of the participant demographic information is presented in Table 1a.

**Table 1.**
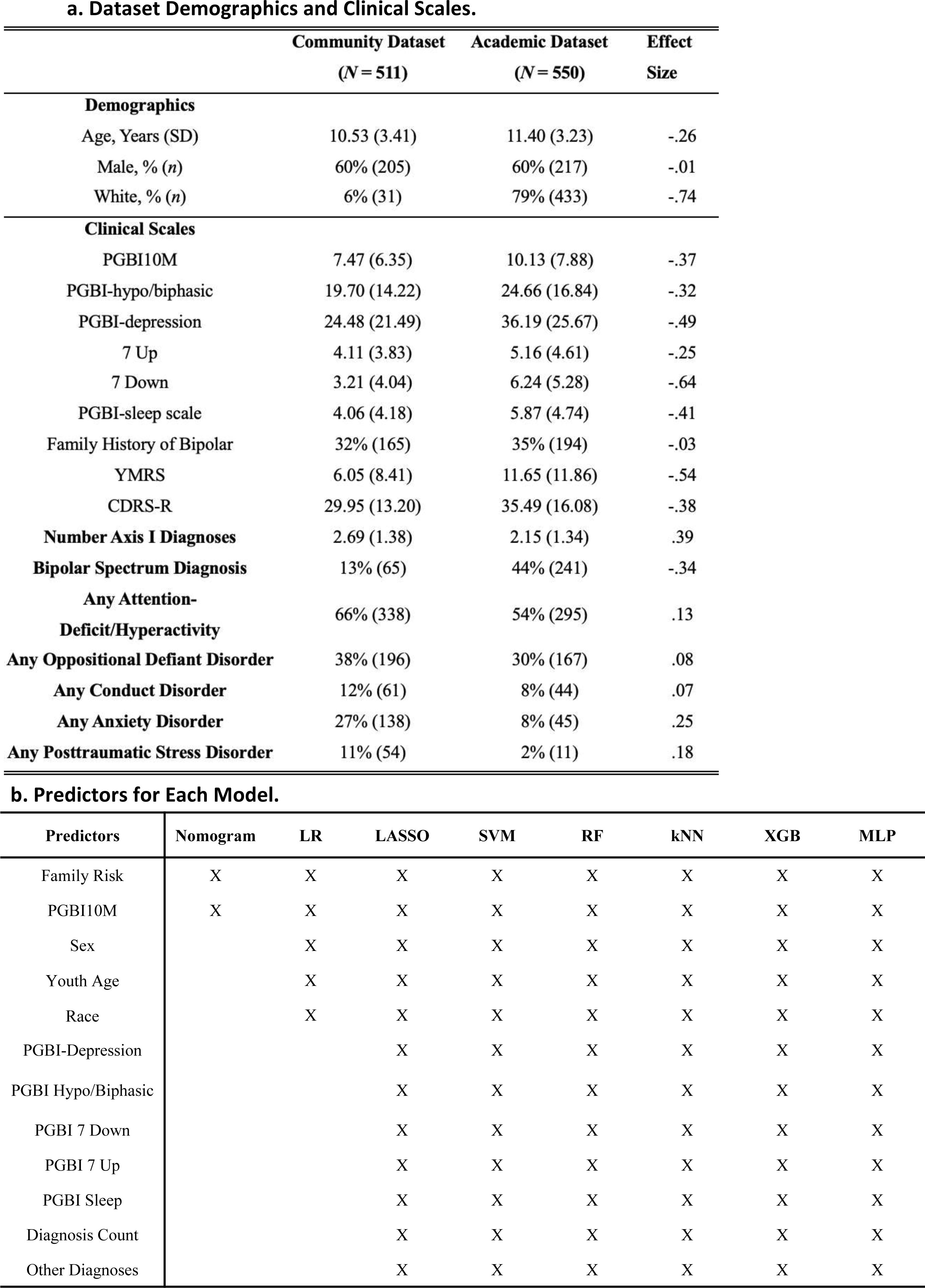
a. Participants Demographics and Clinical Scales. b. Predictors for Each Model.

### Measures and Diagnoses

The Schedule for Affective Disorders and Schizophrenia for School-Age Children–Epidemiologic Version (K-SADS-E) (Orvaschel and Puig-Antich, 1987) or the Present and Lifetime Version (K-SADS-PL) (Kaufman et al., 1997) supplemented with additional mania and depression questions from the Washington University version (Geller et al., 2000) were administered by research assistants to youths and their caregivers. The interrater reliability was kappa of 0.85 or higher at the symptom level. The established diagnoses were derived through a consensus-based review conducted by a qualified psychologist or psychiatrist (Findling et al., 2005; Youngstrom et al., 2005). Diagnostic checklists were masked for diagnoses. Binary coding indicated presence of bipolar spectrum disorder, assigning a “yes” for participants fulfilling the DSM-IV criteria for Bipolar I, Bipolar II, Cyclothymia, or Bipolar Disorder Not Otherwise Specified (Guze, 1995) (renamed Other Specified Bipolar and Related Disorders in later nosology). Diagnoses for the community sample were retrieved from the medical records and assigned the same binary codes. Binary codes were used as categorical labels for classification and to guide supervised learning in machine learning and deep learning models.

We included demographic factors (sex, age, race), family history of bipolar disorder, PGBI scales, other diagnoses, and diagnosis count as predictors in the models. Each model’s choice of predictors is reflected in Table 1b. Demographic factors were recorded from interviews with children and caregivers. Caregivers also reported family history of bipolar disorder, which was binary coded as a yes/no variable. Caregivers completed the Parent General Behavior Inventory (PGBI) about the youth.

The comprehensive version of PGBI encompasses 73 items, each scored on a scale from 0 to 3 (Youngstrom et al., 2001). Because of the PGBI’s reading level requirement and extensive length, various abbreviated versions have been developed. The 10-item mania scale (PGBI-10M) was designed to concentrate on items that most effectively differentiate bipolar from non-bipolar diagnoses through parental reports (Youngstrom et al., 2008). This scale has consistently been recognized for its exceptional discriminative validity among existing checklists (Youngstrom et al., 2015). The abbreviated version comprised of seven items focusing on sleep disturbances also demonstrated high internal consistency and efficacy in identifying mood disorder cases (Meyers and Youngstrom, 2008). Another pair of scales, the 7 Up-7 Down, encompassing seven items for hypomanic/biphasic and depressive symptoms, was also conducted in a self-report format (Youngstrom et al., 2013). The scoring protocol for these scales enables the calculation of fractional scores, prorating scores if respondents skipped an item. In our study, the PGBI-10M and family risk were used in all models according to their importance in clinical diagnosis (Youngstrom et al., 2008; Youngstrom et al., 2008).

### Models

We evaluated a spectrum of predictive models spanning clinical decision tools, statistical models, machine learning, and deep learning models (Figure 1). This design allowed us to systematically compare model performance across increasing levels of complexity.

**Figure 1.**
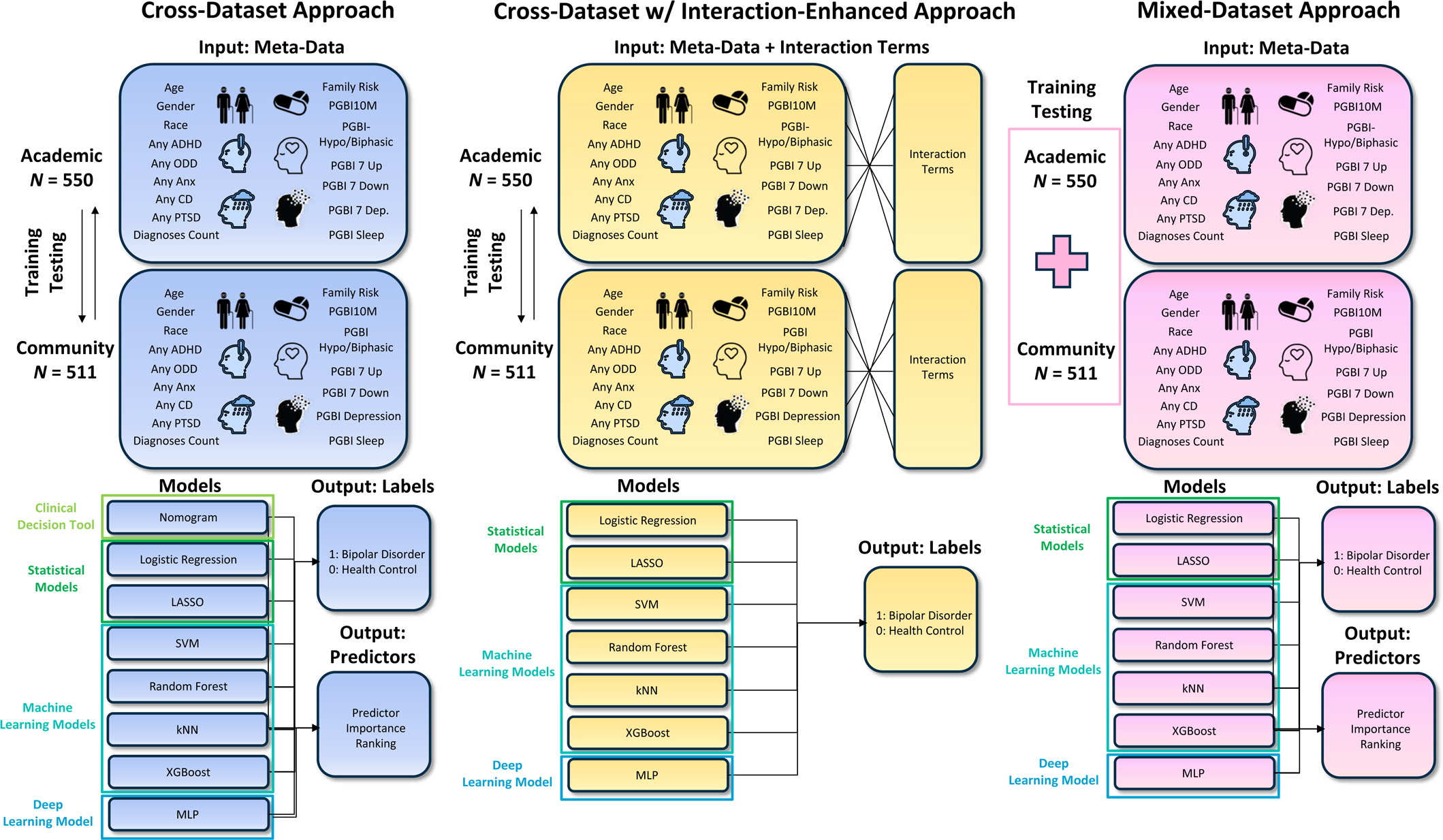
Overview of study design and modeling framework. Predictor variables included demographic factors, family risk, PGBI scales, comorbid diagnoses, and the number of diagnose. Three modeling strategies were compared: baseline cross-dataset, interaction-enhanced cross-dataset, and mixed-dataset approaches. Models ranged from clinical decision tools to statistical, machine learning, and deep learning methods, all performing binary classification. Predictor importance rankings were evaluated across models and approaches.

A nomogram as a clinical decision tool was implemented based on PGBI-10M and family risk. Nomograms are evidence-based graphical tools for estimating individualized risk and supporting clinical decision-making (Straus et al., 2018). We selected this tool as a clinically interpretable benchmark that reflects established diagnostic best practices in pediatric bipolar disorder and provides a reference for evaluating more complex models. Based on the rate of teaching and adoption over several decades (Jaeschke et al., 1994; Youngstrom et al., 2020), as well as changes in the healthcare workforce in the United States (with an increasing reliance on masters level providers, with concomitant abbreviation of diagnostic training) (Youngstrom, 2026), the nomogram may be a reasonable “upper bound” estimate of what a multidisciplinary workforce could implement without shifting to a more computing-intensive hybrid model.

Statistical models included logistic regression (LR) and least absolute shrinkage and selection operator (LASSO). Logistic regression is a widely used probabilistic classification model that estimates the relationship between predictors and binary outcomes (Domínguez-Almendros et al., 2011). We included logistic regression as an interpretable baseline linear model for comparison. LASSO extends linear models by introducing L1 regularization to perform variable selection and improve model stability in high-dimensional settings (Tibshirani, 1996). We selected LASSO to evaluate whether regularization-based feature selection improves generalizability.

Machine learning models included support vector machines (SVM), random forests (RF), k-nearest neighbors (kNN), and extreme gradient boosting (XGBoost). SVM is a margin-based classifier that identifies an optimal linear boundary in feature space (Cortes and Vapnik, 1995), and was selected for its robustness in high-dimensional data. RF is an ensemble learning method that constructs multiple decision trees and aggregates their predictions to reduce variance and improve stability (Breiman, 2001), and was included for its strong performance and interpretability via feature importance ranking. kNN is a non-parametric method that classifies new datasets based on similarity to the neighboring cases (Cover and Hart, 1967), and was included as an approach mirroring real-world clinicians’ experience-based diagnosis. XGBoost is a gradient boosting framework that sequentially builds decision trees to optimize predictive performance (Chen and Guestrin, 2016), and was selected for its ability to capture complex nonlinear relationships.

A deep learning model was also implemented using a multilayer perceptron (MLP), a feedforward neural network capable of modeling complex nonlinear relationships through layered representations (Murtagh, 1991; Rumelhart et al., 1986). We selected MLP to assess whether modern deep learning models improve predictive performance. To mitigate overfitting, relatively simple network architectures with regularization were used.

Detailed model specifications and parameters are provided in the Supplemental Materials. The predictors for each model are shown in Table 1b. Nomogram used two key predictors, LR utilized five predictors, and other models used all predictors listed.

### Modeling Approaches

We compared three training and modeling strategies to evaluate model generalizability across clinical settings, as shown in Figure 1.

In the cross-dataset approach, models were trained on the academic dataset and directly evaluated on the community dataset. This approach simulates real-world deployment of a model developed in the academic setting and applied to the community clinic. The pipeline was reversed to train on the community dataset and evaluate on the academic dataset for further tests of generalization ability. Hyperparameters were tuned using 10-fold cross-validation within the training dataset. For example, LASSO used cross-validated regularization parameters, SVM tuned cost parameters, RF and kNN were optimized using grid search with cross-validation, XGBoost used cross-validation with early stopping to determine the optimal number of boosting iterations, and MLP was tuned using grid search over network size and regularization parameters with cross-validation.

In the cross-dataset with interaction-enhanced approach, two-way interaction terms among predictors were included in LASSO, SVM, RF, kNN, XGBoost, and MLP during model training to capture higher-order relationships. Only two-way interaction terms were considered, as more complicated interactions substantially increase model complexity, are difficult to interpret clinically, and may lead to overfitting and unstable estimations. The same cross-dataset evaluation framework and tuning procedures were applied. This approach was designed to examine whether increasing model variables through interaction terms improves generalizability.

In the mixed-dataset approach, the academic and community datasets were combined into a pooled dataset. The pooled sample was then randomly split into training (70%) and testing (30%) sets using stratified sampling based on the outcome variable. This approach allows models to learn from a more diverse set of clinical presentations and aims to improve robustness under dataset heterogeneity. The same cross-validation strategies were used for hyperparameter tuning.

### Model Evaluations

Model performance was evaluated based on discrimination, calibration, and predictor importance ranking.

Discrimination was assessed using the area under the receiver operating characteristic curve (AUC). AUC quantifies a model’s ability to correctly separate individuals into classes and is widely used for evaluating classification performance. Confidence intervals and standard errors for AUC were estimated to assess statistical uncertainty.

Calibration was evaluated to assess the agreement between predicted probabilities and observed outcomes. We used multiple complementary metrics, including Spiegelhalter’s *z*-test to assess calibration statistically, calibration plots to visualize agreement between predicted and observed risk, Brier scores to quantify overall prediction error, and Nagelkerke’s *R^2^* to measure explained variation. Calibration was evaluated in both internal and external validation settings. In addition, logistic recalibration (Steyerberg, 2009; Van Calster et al., 2019) was applied in the cross-dataset approach internal and external validations to examine whether recalibration improves model generalizability.

Predictor importance consistency was examined across models and approaches. For each model, predictor importance was derived using model-specific approaches, including regression coefficients for logistic regression, non-zero coefficients for LASSO, weight vectors for SVM, feature importance measures for RF and XGBoost, and permutation-based importance for MLP. Predictors were ranked by their contribution to model performance, and the top predictors from each model were recorded. These predictors were then counted across models to compute selection frequency and identify consistently important variables across modeling strategies and clinical settings.

## Results

### Baseline Cross-Dataset Models Performance

In the baseline cross-dataset setting, all models demonstrated good discrimination ability in internal validation within the academic dataset, with AUC values ranging from approximately 0.88-0.93 (Figure 2a and Figure 2c). Machine learning and deep learning models achieved the highest performance, including RF (AUC ≈ 0.93), XGBoost (AUC ≈ 0.93), and MLP (AUC ≈ 0.92). Notably, the nomogram and logistic regression models showed comparable performance with an AUC of 0.89 for both.

**Figure 2.**
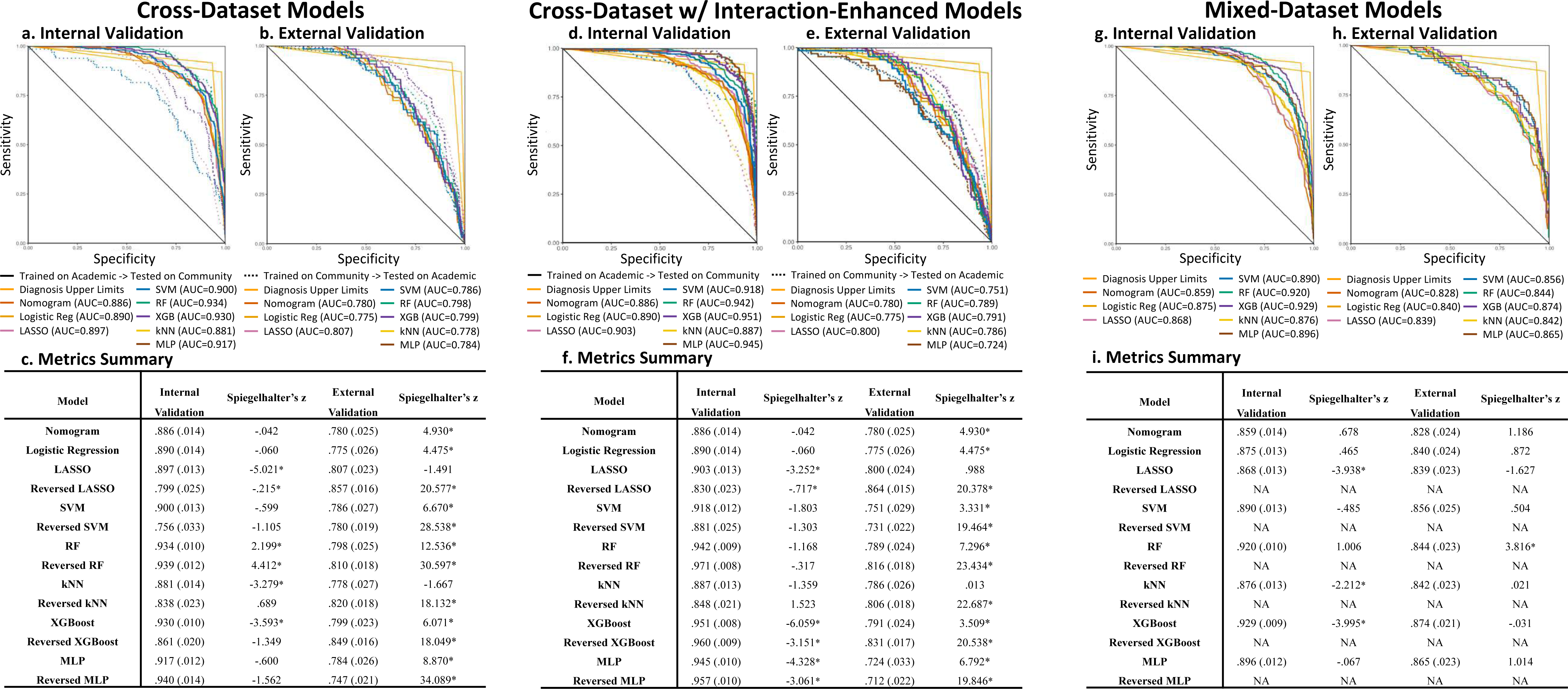
Model discrimination and calibration across modeling strategies. ROC curves and performance summaries are shown for baseline cross-dataset models (a-c), interaction-enhanced models (d-f), and mixed-dataset models (g-i). Panels (a), (d), and (g) display internal validation performance, while panels (b), (e), and (h) show external validation results. Panels (c), (f), and (i) summarize discrimination (AUC) and calibration (Spiegelhalter’s *z*-test) metrics across models. Solid line models were trained on the academic sample and externally validated on the community sample, whereas dashed-line models were trained on the community sample and externally validated on the academic sample.

However, when the trained models were applied to the community dataset for external validation, model performance decreased substantially across all methods, with AUC values declining to approximately 0.75-0.81 (Figure 2b-c). In addition, performance differences between models became less distinct, and model performances became similar regardless of complexity. Calibration analysis further revealed instability across models, with significant Spiegelhalter’s z-values observed in either internal or external validation for all models.

In the reverse validation setting, where models are trained with the community dataset and tested on the academic dataset, a similar pattern was observed (Figure 2a-c). Internal validation performance in the community dataset showed a slightly wider range with AUC **≈** 0.76-0.94, while external validation performance on the academic dataset was slightly higher (AUC ≈ 0.78-0.86). However, calibration instability was more prominent, with all reversed models exhibiting significant Spiegelhalter’s *z*-test values in external validation.

To further examine calibration performance under cross-dataset settings, we evaluated baseline models trained on the academic dataset and tested on the community dataset before and after logistic recalibration (Figure 3). In internal validation within the academic dataset before recalibration, calibration curves for most models were generally close to the diagonal, suggesting reasonable agreement between predicted and observed probabilities (Figure 3, left panel). However, several models, including LASSO, RF, kNN, and XGBoost, showed significant Spiegelhalter’s *z*-test values (−5.0 to 2.2), indicating residual miscalibration despite visually acceptable calibration. However, external validation revealed systematic miscalibration across all models. Calibration curves consistently deviated below the diagonal (Figure 3, left panel, second column), indicating that predicted probabilities were generally higher than observed outcomes, particularly in the medium-to-high risk region around 0.50 in observed probability. Spiegelhalter’s *z*-values ranged approximately from −0.06 to 5.00, with corresponding significant *p*-values across models. Brier scores were also elevated (0.17-0.21), reflecting increased prediction error. Miscalibration was more pronounced in more complex models, including SVM, RF, XGBoost, and MLP, which exhibited larger deviations from the ideal calibration line.

**Figure 3.**
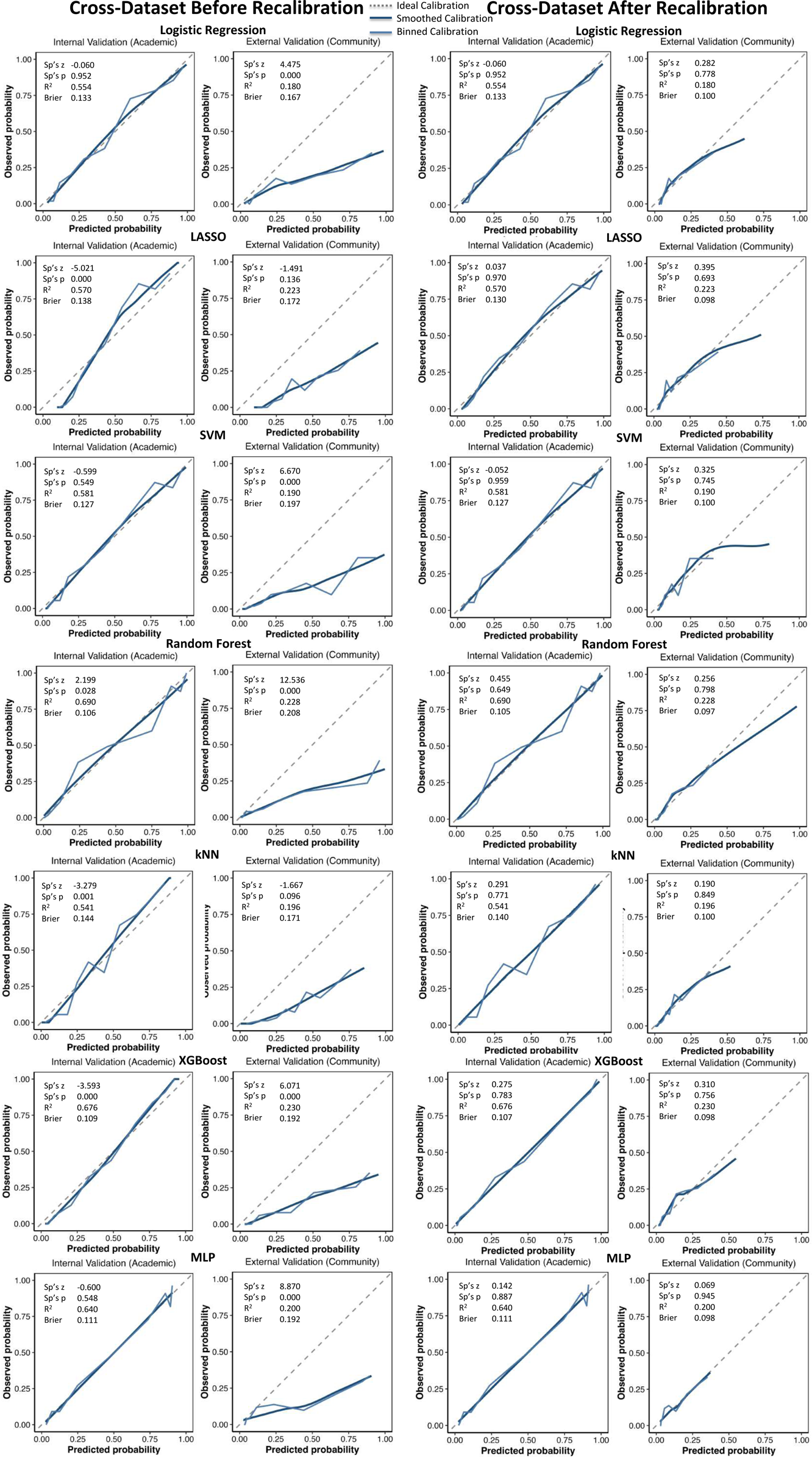
Calibration performance of baseline models with and without recalibration. Calibration plots for baseline cross-dataset models are shown before (left panel) and after (right panel) recalibration across internal (academic) and external (community) validation settings. The dashed diagonal line represents ideal calibration. The solid dark blue lines indicate smoothed model calibration curves, while the solid light blue lines represent binned calibration curves.

After applying logistic recalibration, calibration improved substantially across both internal and external validation (Fig. 3, right panel). In internal validation, calibration metrics stabilized across all models, with Spiegelhalter’s *z*-values falling below 0.50 and all *p*-values non-significant (*p* > 0.60). Brier scores decreased for several models, including LASSO, RF, kNN, and XGBoost, while *R^2^* values remained unchanged, indicating that recalibration primarily adjusted probability scaling without altering discrimination. In external validation, recalibration led to a prominent improvement in calibration performance. Calibration curves were notably closer to the diagonal across all models, and Brier scores decreased substantially to ≤ 0.10. Spiegelhalter’s *z*-values were reduced to a narrow range of 0.07 to 0.40, with corresponding non-significant *p*-values (*p* > 0.65), indicating well-calibrated predictions. Although slight overestimation persisted in the high-risk region, overall agreement between predicted and observed probabilities showed considerable improvement.

### Interaction-Enhanced Cross-Dataset Models Performance

Incorporating two-way interaction terms slightly improved internal validation performance across all models (Figure 2d and Figure 2f), with discrimination ability ranging from approximately 0.89 to 0.95. However, these improvements did not translate to external validation. When externally evaluated on the community dataset, most models performed slightly worse than baseline models, with AUC values ranging from approximately 0.72 to 0.80 (Figure 2e), consistent with over-fitting the training data. The only exception was kNN, which maintained comparable or slightly improved performance, likely due to its instance-based nature and ability to leverage local similarity across cases. Calibration remained unstable across models, with most approaches exhibiting significant Spiegelhalter’s *z*-test values in either internal or external validation.

### Mixed-Dataset Models Performance

Models trained on the mixed-dataset demonstrated strong performance in both internal and external validation (Figure 2g-i). Internal validation AUC values were high across all models, ranging from 0.86 to 0.93, comparable to those observed in the baseline setting. External validation performance showed substantially improved consistency with internal validation, with AUC values ranging from approximately 0.83 to 0.87 (Fig. 2h). Unlike the cross-dataset approaches, performance degradation in the mixed-dataset external validation was minimal. Calibration metrics further indicated improved stability, with much smaller and less frequently significant Spiegelhalter’s *z*-test values across models (Fig. 2i).

To further examine calibration performance under the mixed-dataset setting, we evaluated calibration curves for all models in both internal and external validation (Figure 4). Overall, calibration performance without recalibration was substantially improved compared to cross-dataset approaches with recalibration. The calibration curves closely align with the ideal diagonal line across most models. In internal validation, all models except LASSO, kNN, and XGBoost showed Spiegelhalter’s *z*-values ≤ 1.0 and non-significant *p*-values. Brier scores were consistently low across models (all < 0.15), suggesting good overall prediction accuracy. In external validation, calibration performance remained stable and closely matched internal validation. Calibration curves were generally close to the diagonal, indicating strong agreement between predicted and observed probabilities. Brier scores remained low across all models (≤ 0.15). Spiegelhalter’s *z*-values ranged from approximately −1.63 to 0.02, with non-significant p-values across all models except RF, indicating overall well-calibrated predictions. *R^2^* values were consistently higher than those observed in cross-dataset approaches in Figure 3, suggesting improved reliability of probability estimation when models were trained on mixed datasets.

**Figure 4:**
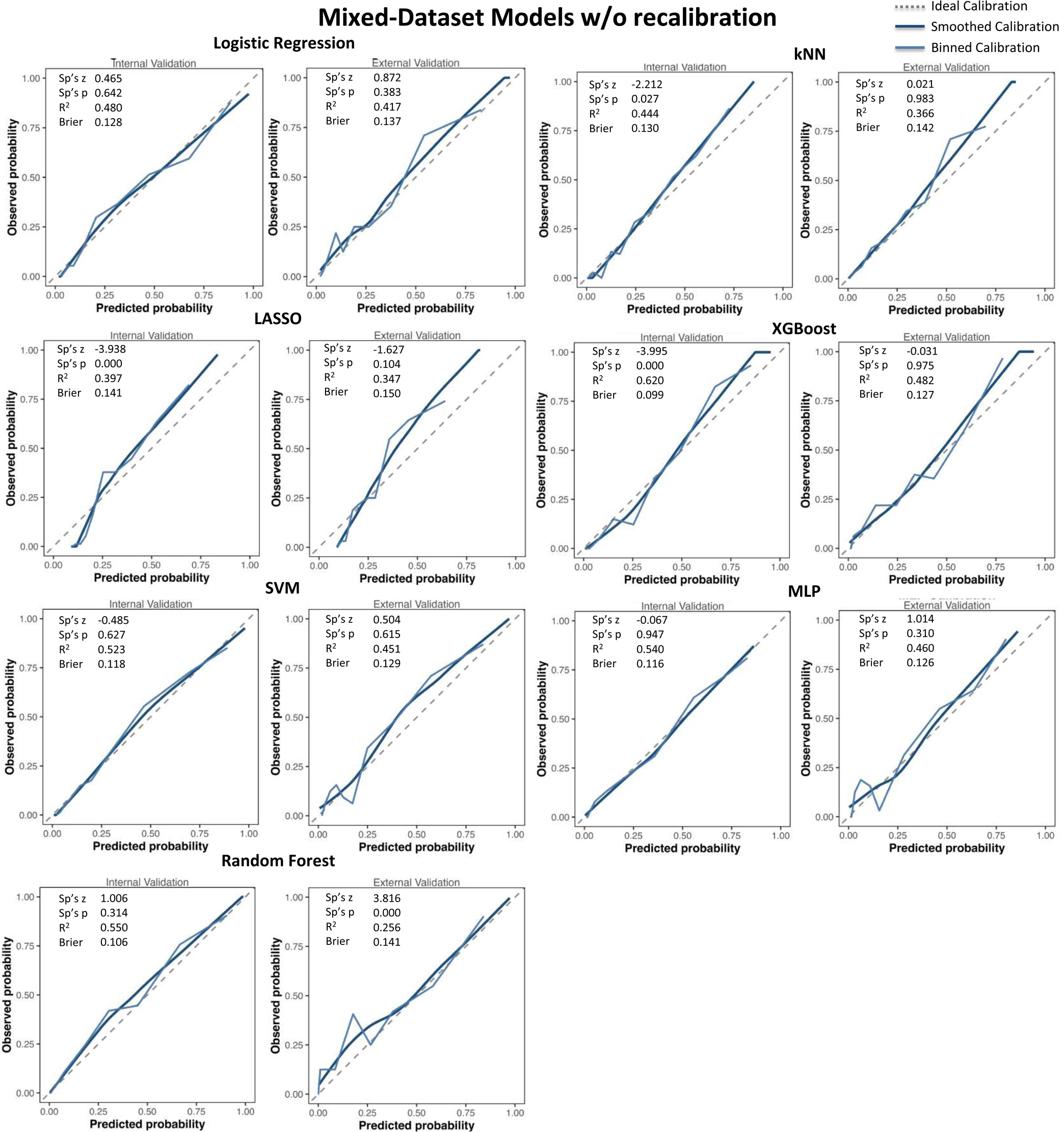
Calibration performance of mixed-dataset models. Calibration curves for models trained on the pooled academic and community datasets and evaluated in internal and external validation. The dashed diagonal line represents ideal calibration. The solid dark blue lines indicate smoothed model calibration curves, while the solid light blue lines represent binned calibration curves.

### Predictor Importance Ranking

Predictor importance rankings were examined across models to identify variables that consistently contributed to prediction performance (Table 2). In the baseline cross-dataset models (Table 2a), family risk and PGBI-10M were consistently identified as the most important predictors across all models (6 out of 6 models), indicating robust and stable contributions to model performance regardless of modeling approach. In addition to these core predictors, several other variables were frequently selected, including the number of diagnoses, additional PGBI symptom scales, and other psychiatric diagnoses. These predictors were less consistently identified in simpler statistical models but were more prominent in machine learning and deep learning models. In the mixed-dataset models (Table 2b), the overall pattern of predictor importance remained largely consistent. Family risk and PGBI-10M continued to be the dominant predictors across all models (6 out of 6 models), further supporting their stability across different training strategies and data distributions. Race emerged as an important variable in all models when trained on the mixed dataset. Other predictors, including the number of diagnoses, additional PGBI symptom scales, and the age of the child, were also frequently identified, primarily by machine learning and deep learning models.

**Table 2.**
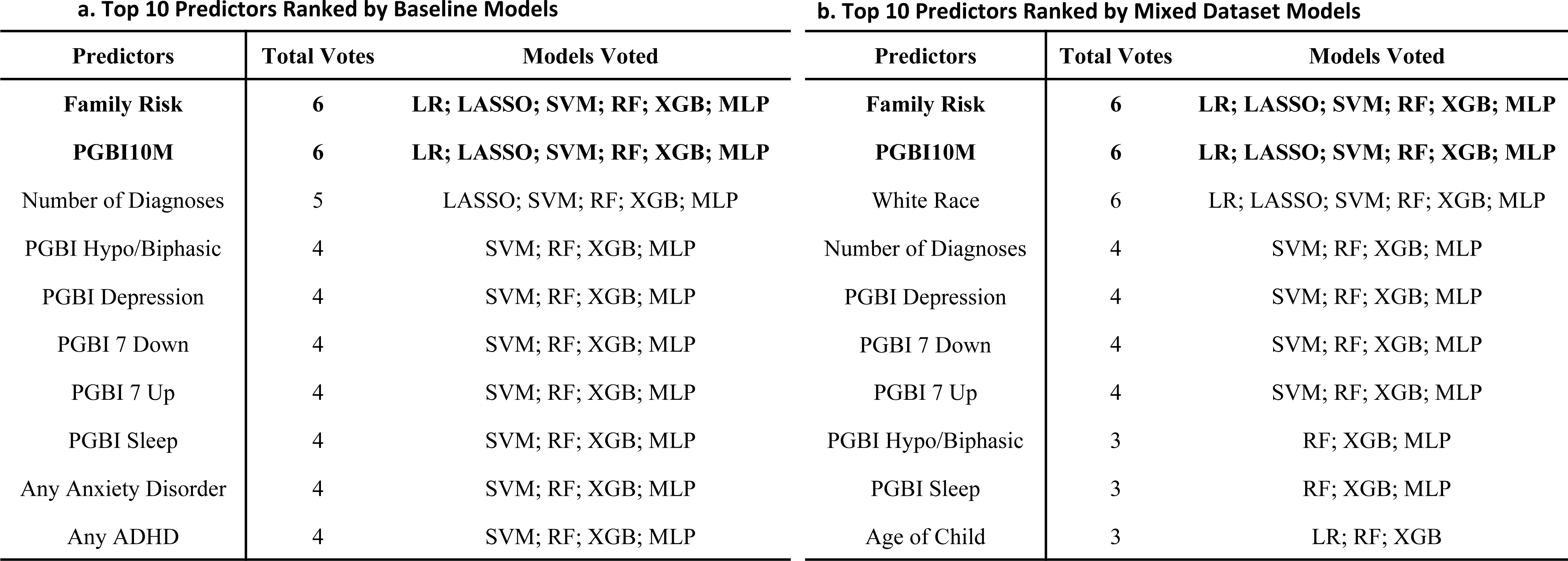
Predictor importance rankings. a. predictor importance ranking with baseline cross-dataset models. b. predictor importance ranking with mixed-dataset models.

## Discussion

We systematically evaluated the performance and generalizability of multiple prediction models for pediatric bipolar disorder across different clinical settings. While all models demonstrated strong discrimination in internal validation, their performance shrank substantially under cross-dataset external validation, with calibration deteriorating more than classification accuracy. These findings highlight significant challenges in model transportability across clinical contexts. Notably, increasing model complexity did not translate into improved external performance and was associated with greater miscalibration. In contrast, training on pooled, multi-site datasets prominently improved both discrimination and calibration, underscoring the pivotal role of data diversity in developing robust and clinically applicable prediction models. Furthermore, recalibration proved to be a simple yet effective strategy to restore probability accuracy across settings, suggesting that miscalibration is largely driven by differences in probability scaling rather than by fundamental changes in the relationships between predictors and the diagnosis. Across all training approaches and models, key predictors such as family risk and PGBI-10M demonstrated consistent importance, supporting the presence of stable and clinically meaningful signals underlying bipolar disorder risk. The emergence of sociodemographic variables in mixed datasets highlights the importance of contextual factors when applying predictive models across diverse populations.

Data diversity, rather than model complexity, appeared to be the primary determinant of generalizability in psychiatric prediction models. Although more complex machine learning and deep learning models are often assumed to provide superior predictive performance (Pigoni et al., 2024), our results indicate that such advantages do not extend to external validation. In fact, more complex models tended to exhibit greater miscalibration when transported to a different clinical setting, suggesting increased sensitivity to dataset-specific patterns (Van Calster et al., 2019). This finding is particularly relevant for clinically heterogeneous conditions such as pediatric bipolar disorder, where symptom presentation varies widely across individuals and settings (Kowatch et al., 2005; Van Meter et al., 2016). Models trained on relatively homogeneous datasets may fail to capture this variability, limiting their ability to generalize. Incorporating more diverse, multi-site data appears to improve model robustness by reflecting the range of real-world clinical presentations.

Consistent with prior work on dataset shift (Beam and Kohane, 2018; Konig et al., 2007; Subbaswamy and Saria, 2020), we observed substantial degradation in model performance when applied across clinical settings. Differences in patient populations, referral patterns, and clinical severity likely contributed to this effect. While discrimination metrics reflect a model’s ability to predict risk, calibration determines whether predicted probabilities correspond to actual risk, which is critical for clinical decision-making. Poor calibration may lead to biased clinical decisions even when discrimination is acceptable (Alba et al., 2017; Lindhiem et al., 2018). In our study, models applied across datasets tended to overestimate risk, which could translate into unnecessary clinical concern or potential overdiagnosis in practice. Moreover, adding interaction terms provided modest improvements in internal performance but did not improve external generalizability, in fact worsening calibration. These interaction effects may capture dataset-specific relationships that do not generalize across populations, thereby increasing the risk of overfitting and capturing associations that may not reflect true underlying clinical relationships.

Recalibration is a practical and effective approach to address cross-site miscalibration. By adjusting predicted probabilities without altering the underlying model structure, recalibration substantially improved agreement between predicted and observed outcomes while preserving discrimination ability, consistent with prior work demonstrating that recalibration primarily corrects probability scaling without altering model performance (Janssen et al., 2008). This finding suggests that transportability issues are often driven by differences in probability scaling rather than changes in the underlying associations between predictors and outcomes. From a clinical perspective, recalibration offers a feasible strategy for adapting existing models to new settings without full model retraining, which may be helpful in resource-limited environments. The probability nomogram method can use local base rates as the prior probability estimate, implementing a simple and feasible recalibration method.

Our findings highlight the advantages of pooled, multi-site training. Models trained on mixed datasets achieved improved performance in both discrimination and calibration without recalibration, suggesting enhanced transportability and generalization. This improvement reflects that exposure to a broader range of symptom presentations and clinical contexts can reduce overfitting to site-specific patterns (Debray et al., 2019; Subbaswamy and Saria, 2020). These results support the growing emphasis on multi-site data integration in psychiatric research and underscore the importance of open and collaborative datasets for improving model robustness and increasing clinical applicability. Research coalitions such as the National Network of Depression Collaborative, PEDSNet, CAPTN, and other learning networks may be particularly valuable by creating opportunities for mixed-data applications, accomplishing diversity in geographical, demographic, and referral patterns. At the same time, present results also underscore the importance of having high-validity inputs, such as the PGBI-10M or a pragmatic measure of family history, as these were consistently major contributors to all successful models.

We observed high consistency in the importance of predictors across training approaches and all models. The most commonly used clinical variables, including family risk and PGBI-10M (Youngstrom et al., 2008; Youngstrom et al., 2008), were ranked as the most important predictors by all models that could show the predictor ranking, suggesting robust underlying clinical signals. Compared to clinical decision tools, the statistical methods, machine learning, and deep learning models were able to incorporate a broader range of predictors and offer complementary insights into variable importance. Among these models, machine learning and deep learning models are more sensitive at detecting important predictors than statistical models, and thus may serve as useful tools for identifying complex predictor patterns and supporting clinical decision-making (Cearns et al., 2019; Quinn et al., 2024). Notably, sociodemographic variables such as race and age emerged as important predictors in mixed datasets (DelBello et al., 2001; Jenkins and Youngstrom, 2016), which may reflect differences in access to care, diagnostic practices, or broader population characteristics (Youngstrom, 2026). These findings highlight the importance of considerations regarding potential bias in model deployment and health disparities. However, only some of the models allow recovery of predictors, limiting the theory-building value of some models.

Although our findings offer important insights into generalizable predictive modeling for pediatric bipolar disorder, several limitations should be considered. First, as in many clinical prediction studies, the outcome labels were derived from clinician-based diagnostic assessments rather than an objective gold standard. Both data sets used a longitudinal expert evaluation of all available data (LEAD) standard, considered best practice in psychiatry (Spitzer, 1983), and interviewers and consensus experts were shared across both settings for the research diagnoses. Although these diagnoses reflect best practice, potential misclassification still may have been introduced into both model training and evaluation, which could affect model performance estimates (Kraemer, 1992). We estimated the effect of the reliability of the LEAD consensus diagnoses, setting potential ceiling on classification accuracy (shown as the diagnosis AUC in the discrimination figures). Second, the set of predictors was pre-specified based on available clinical variables, which may limit the identification of additional relevant features or novel predictors. Future studies incorporating broader and multiple data sources may exhibit different predictor preferences. Conversely, the robust performance of the PGBI and family history attest to the value of including predictors that leverage prior empirical work (Alcaíno et al., 2024; Birmaher et al., 2010; Hodgins et al., 2002). Third, while machine learning and deep learning models offer flexibility in capturing complex relationships, their limited interpretability may pose challenges for clinical adoption. Approaches such as examining predictor importance can provide some insights into model behavior, but they may not fully address concerns regarding transparency in clinical decision-making.

### Conclusions

Although predictive models for pediatric bipolar disorder achieved strong internal performance, their generalizability across clinical settings was limited, primarily due to dataset shift and miscalibration, and to a lesser extent, instability in choice of second-tier predictors. Recalibration improved probability estimates without altering discrimination, suggesting that transportability issues are largely related to probability scaling. Increasing model complexity did not improve external performance, whereas training on pooled datasets substantially enhanced both discrimination and calibration, suggesting the importance of open and collaborative datasets. First-tier predictors such as family risk and PGBI10M remained consistent across models, supporting the robustness of the underlying clinical signals and showing the benefit of including predictors with a strong empirical evidence base. These findings highlight that data diversity, rather than model complexity, is critical to developing clinically useful and generalizable prediction models in complex psychiatric disorders like pediatric bipolar disorder.

## Supporting information

Supplemental Model Specifications and Parameters

## Data Availability

All data produced in the present study are available upon reasonable request to the corresponding author

## Authorship Contribution Statement

Zhuoyu Shi selected the machine learning and deep learning models, as well as proposing the mixed-data methodology for pooling samples, conducted the analyses and made the figures, and drafted the manuscript. Dr. Eric Youngstrom supervised the project and revised the manuscript. Yinuo Liu revised the manuscript. Dr. Eric Youngstrom and Yinuo Liu reviewed the statistical analyses and summary figures. Dr. Jennifer Youngstrom supervised data collection and reviewed the manuscript. Dr. Robert Findling contributed to data collection, offered advice for the project, and reviewed the manuscript.

