## Supplemental Model Specifications and Parameters for "Data Diversity vs. Model Complexity in the Prediction of Pediatric Bipolar Disorder: Evidence from Academic and Community Clinical Samples"

All models were implemented in R (version 2024.12.1). Additional details regarding model implementation are available from the corresponding author and the first author upon request.

**Nomogram**

Nomograms are graphical representations of regression-based prediction models that estimate individualized risk probabilities and are commonly used in clinical decision-making because of their simplicity and transparency. In this study, we calculated the probability based on PGBI-10M and the binary input of family bipolar disorder.

**Logistic Regression**

Logistic regression was included as the primary statistical baseline because it is one of the most widely used and interpretable methods for binary classification in clinical research. It estimates the log-odds of the outcome as a linear combination of predictors, generating directly interpretable coefficients and predicted probabilities.

In the cross-dataset, no-interaction approach, the final academic logistic regression model was trained using five predictors: PGBI-10M, family risk, sex, age, and race. The final model was fit with a binomial logit link using the glm function in R. The estimated coefficients in the academic training sample were: intercept = -2.74, PGBI-10M = 0.21, family risk = 1.83, sex = -0.2958, age = -0.0484, and race = 0.4284.

**LASSO**

LASSO adds an L1 penalty to the logistic regression objective function, shrinking some coefficients toward zero and allowing automatic feature selection. For LASSO, hyperparameters were optimized using 10-fold cross-validation, with AUC as the tuning criterion. Predicted probabilities for both internal and external evaluation were generated using the lambda.1se solution, which favors a more conservative and parsimonious model than lambda.min.

**Support Vector Machine (SVM)**

Support vector machine (SVM) is a machine learning classifier capable of identifying a separating hyperplane that maximizes the margin between classes. SVM is particularly useful in settings with multiple correlated predictors.

In this study, SVM models were implemented using a linear kernel to balance predictive flexibility and interpretability. Models were trained using tune.svm with C-classification and probability estimation enabled (probability = TRUE). Hyperparameters were tuned using 10-fold cross-validation over a cost grid of 0.001, 0.005, 0.01, 0.05, and 0.1. For the academic-trained model, the optimal cost parameter was 0.01. For the community-trained model, the optimal cost parameter was 0.001.

**Random Forest (RF)**

Random forest is an ensemble learning model that aggregates predictions from multiple decision trees. This approach is often robust to noise, can capture nonlinear effects, and provides variable importance measures.

RF models were fit using the caret::train framework with the rf method. Hyperparameters were tuned using 10-fold cross-validation with ROC as the optimization metric. The tuning grid evaluated included mtry = 2, 3, 4, 5, and 6. The number of trees was fixed at 500 (ntree = 500), and tree complexity was constrained using maxnodes = 15 and nodesize = 15 to reduce overfitting. In the academic-trained model, the optimal tuning value was mtry = 6. In the community-trained model, the optimal tuning value was mtry = 3.

**k-Nearest Neighbors (kNN)**

k-Nearest Neighbors is a non-parametric, distance-based machine learning classifier. It predicts the class of a new case based on the class of its nearest neighbors in the training set, providing a simple but informative approach for comparison with more structured and ensemble-based methods.

Because kNN is sensitive to feature scale, all predictors were centered and scaled prior to model fitting. Models were trained using the caret::train framework with 10-fold cross-validation and ROC as the optimization metric. The tuning grid evaluated included k = 5, 7, 9, 11, 15, 21, 31, and 41. For both the academic-trained and community-trained models, the optimal tuning value was k = 41.

**Extreme Gradient Boosting (XGBoost)**

XGBoost is a boosting-based ensemble method that sequentially builds shallow decision trees to improve predictions by correcting errors from previous trees. Compared with bagging-based ensemble methods such as RF, XGBoost can be more flexible and efficient in modeling complex nonlinear relationships.

In this study, XGBoost was implemented using the xgboost package with a binary logistic objective and AUC as the evaluation metric. Predictor matrices were converted to numeric matrices and stored as xgb.DMatrix objects. A conservative parameter setting was used to limit overfitting: max_depth = 2; eta = 0.03; subsample = 0.7; colsample_bytree = 0.7; min_child_weight = 3; gamma = 0.5. The maximum number of boosting rounds was set to 1000, and hyperparameter selection focused on identifying the optimal number of rounds using 10-fold stratified cross-validation with early stopping after 30 rounds without improvement (early_stopping_rounds = 30). The optimal number of rounds was 167 for the academic-trained model and 37 for the community-trained model. Final XGBoost models were then trained using these dataset-specific optimal boosting rounds.

**Multilayer Perceptron (MLP)**

A multilayer perceptron (MLP) is a feedforward neural network that models nonlinear relationships through one or more hidden layers and learned weighted connections between predictors and outcomes. Compared with other models, MLP allows greater flexibility in capturing complex predictor interactions and nonlinear patterns.

Given the risk of overfitting, relatively small network architectures with explicit regularization were used. MLP models were trained using the caret::train interface with the nnet method. Predictors were centered and scaled before training. Hyperparameters were tuned using 10-fold cross-validation with ROC as the optimization metric. The tuning grid included combinations of size = 1, 3, 5 and decay = 0.001, 0.01, 0.1, where size represents the number of hidden units and decay represents the weight decay regularization parameter. The maximum number of iterations was set to 200, and training traces were suppressed (trace = FALSE). For the academic-trained model, the optimal values were size = 1 and decay = 0.1. For the community-trained model, the optimal values were size = 3 and decay = 0.1.
